# Therapeutic Alliance and Treatment Outcomes Among Patients with Depression in Benue State

**DOI:** 10.64898/2026.05.19.26353601

**Authors:** Caleb Onah, Chinelo Helen Ogwuche, Bridget Orako Otumala

## Abstract

Depression remains a major public health concern globally, particularly in low-resource settings where access to quality mental health care is limited and treatment outcomes are often suboptimal. In this context, the quality of the clinician–patient relationship has been increasingly recognised as a critical determinant of therapeutic success. This study examined the influence of clinician–patient therapeutic alliance and relational factors on treatment outcomes among patients with depression in Benue State, Nigeria. A cross-sectional correlational design was adopted, involving patients diagnosed with depression and receiving care in selected health facilities. Data were analysed using Structural Equation Modelling (SEM) to test hypothesised relationships among therapeutic alliance, relational factors, and treatment outcomes. The measurement model demonstrated strong psychometric properties, with all factor loadings exceeding 0.60, composite reliability above 0.90, and adequate convergent and discriminant validity. The structural model showed shows indices (χ^2^/df = 2.67, CFI = 0.95, RMSEA = 0.052, SRMR = 0.043). Results revealed that therapeutic alliance significantly predicted treatment outcomes (β = 0.45, p < 0.001), while relational factors also had a significant positive effect (β = 0.39, p < 0.001). Therapeutic alliance further significantly predicted relational factors (β = 0.33, p < 0.001). The model explained 61% of the variance in treatment outcomes. Mediation analysis indicated that relational factors partially mediated the relationship between therapeutic alliance and treatment outcomes, accounting for 29% of the total effect. The study concludes that therapeutic alliance, strengthened through trust, empathy, and collaboration, plays a central role in improving depression outcomes. Strengthening relational competencies in clinical practice is therefore essential for enhancing mental health care delivery in Nigeria.

## Background

Depression remains one of the most significant contributors to global disease burden, affecting individuals’ emotional, cognitive, and social functioning across diverse settings. Contemporary psychiatric and psychological research increasingly emphasises not only the efficacy of treatment modalities but also the relational processes through which such treatments are delivered (Videtta et al., 2025). Over the past decade, the therapeutic alliance has emerged as a robust predictor of treatment outcomes in depressive disorders. Recent empirical studies indicate that stronger alliance ratings are consistently associated with greater reductions in depressive symptomatology (Oladeji et al., 2025), irrespective of treatment modality, whether pharmacological, psychotherapeutic, or combined approaches (Goodwin et al., 2026). In a large-scale telepsychiatry cohort, alliance scores measured early in treatment significantly predicted clinically meaningful improvements in depression outcomes over time, reinforcing the temporal and causal relevance of relational engagement in care delivery (Phillips et al., 2025).

From a relational perspective, the therapeutic alliance is not merely a supportive adjunct to treatment but a lively mechanism of change. It facilitates patient engagement, enhances adherence to treatment plans, and fosters openness in discussing distressing symptoms (Hehlmann et al., 2026; Onah & Oladejo, 2023). In this study, treatment outcome is defined in terms of treatment adherence and reduction in depressive symptoms. Relational factors were conceptualised as trust, empathy, and effective communication within the clinician–patient interaction. Empirical evidence further demonstrates that disruptions in alliance are associated with poorer outcomes, including persistent depressive symptoms and maladaptive trajectories (Tetzlaff et al., 2025).

Findings suggest that patients with stronger and more stable alliances are more likely to exhibit improvements in suicidal ideation, whereas deteriorating alliances correlate with persistent or worsening risk profiles (Melzer et al., 2024). These findings highlight the protective role of relational continuity and trust within therapeutic encounters. Moreover, the predictive value of therapeutic alliance extends across age groups and treatment contexts. In adolescent depression, convergence between clinician and patient perceptions of alliance has been shown to significantly predict treatment outcomes, suggesting that mutual recognition of the relationship quality is crucial for therapeutic success (Cirasola et al., 2025; Hull et al., 2025; Josiah et al., 2019).

Despite this growing body of global evidence, there remains a paucity of context-specific research examining the clinician–patient therapeutic alliance in sub-Saharan Africa, particularly in Nigeria. Mental health systems in regions such as Benue State are characterised by limited resources, high stigma, and disparities in access to care, all of which may influence the formation and maintenance of therapeutic relationships. Cultural beliefs about mental illness, communication styles, and expectations of care further shape the relational dynamics between clinicians and patients. Consequently, the generalisability of findings from high-income settings to Nigerian contexts remains uncertain.

In addition, depression in Nigeria is often underdiagnosed and undertreated, with patients frequently presenting late or discontinuing treatment prematurely. These patterns may, in part, reflect weaknesses in therapeutic engagement and alliance formation. Understanding how relational factors influence treatment adherence and outcomes is therefore critical for improving clinical effectiveness within this context. The relational perspective provides a valuable framework for examining how interpersonal processes—such as empathy, trust (Anderson & Dedrick, 1990), collaboration, and communication— mediate the impact of clinical interventions on patient outcomes (Onah & Gwar, 2025).

Both clinician behaviours (e.g., empathy, attunement, cultural competence) and patient factors (e.g., readiness for change, interpersonal style, severity of symptoms) contribute to alliance quality and its subsequent impact on outcomes (Videtta et al., 2025; Tetzlaff et al., 2025). This reinforces the need for integrative models that situate alliance within broader socio-cultural and clinical contexts. Given these considerations, there is a compelling need to investigate the clinician–patient therapeutic alliance and its relationship with treatment outcomes among patients with depression in Benue State. Such inquiry is particularly relevant in light of ongoing efforts to strengthen mental health services in Nigeria and to promote patient-centered care. By adopting a relational perspective, this study seeks to contribute to a deeper understanding of how therapeutic relationships influence recovery trajectories, thereby informing clinical practice, training, and policy development in the region.

While the association between therapeutic alliance and treatment outcomes is well established in high-income settings, significantly less is known about the mechanisms through which this relationship operates in low-resource and culturally diverse contexts. In particular, the extent to which relational processes such as trust, empathy, and communication function as mediating pathways within sub-Saharan African mental health systems remains underexplored. This study addresses this gap by examining not only the direct association between therapeutic alliance and treatment outcomes, but also the intermediary role of contextually grounded relational factors. In doing so, the study offers a context-sensitive extension of Therapeutic Alliance research within Nigerian clinical settings.

### Objectives of the Study

1. To examine the nature and strength of the clinician–patient therapeutic alliance among patients diagnosed with depression in Benue State.
2. To determine the relationship between clinician–patient therapeutic alliance and treatment outcomes among patients with depression in Benue State.
3. To assess the influence of selected relational factors (such as trust, empathy, and communication) on treatment adherence and symptom improvement among patients with depression.

### Theoretical Framework: Bordin’s Working Alliance Theory (Relational Model)

One of the most relevant and widely applied theories underpinning the clinician–patient therapeutic alliance is Bordin’s Working Alliance Theory, which conceptualises the alliance as a collaborative partnership between clinician and patient, built on three core components: agreement on goals, agreement on tasks, and the development of a personal bond. Contemporary literature continues to affirm the relevance of this theory, particularly in understanding treatment outcomes in depression (Videtta et al., 2025).

**Diagram 2:**
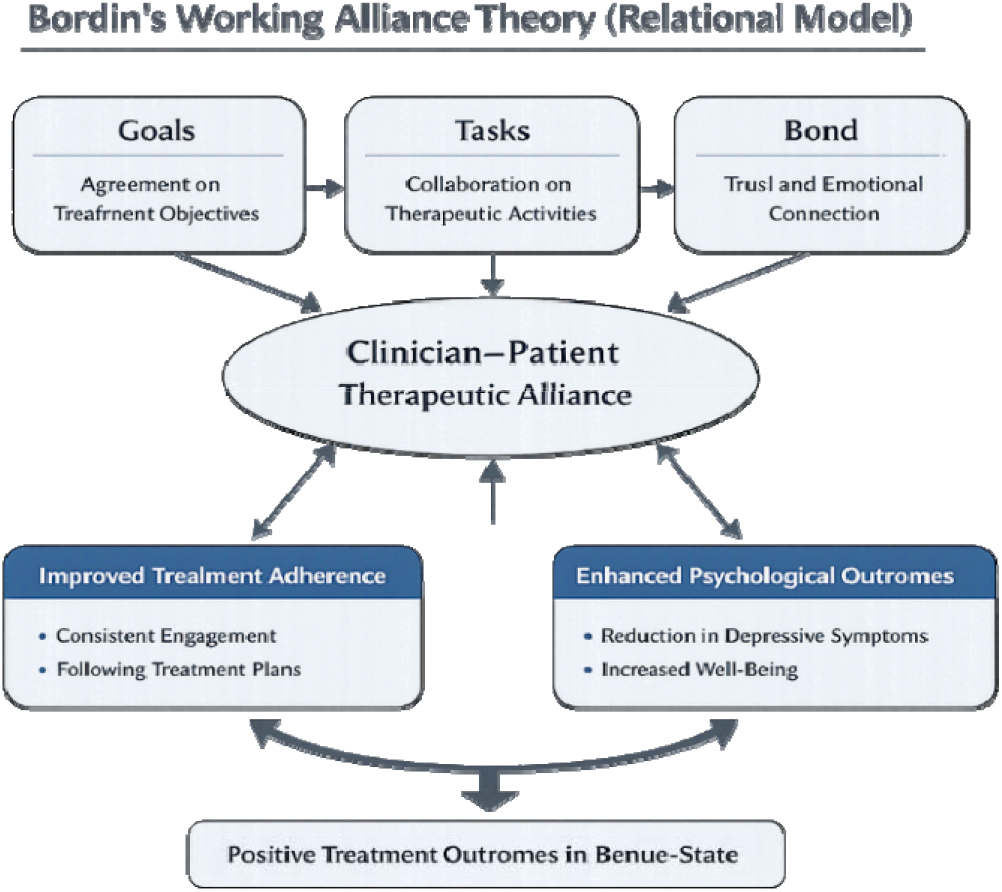
Bordin’s Working Alliance Theory (Relational Model): Application to Clinician–Patient Therapeutic Alliance and Treatment Outcomes in Depression Care.

From a relational perspective, Bordin’s theory posits that the therapeutic alliance is not merely a supportive element but a central mechanism of change in psychotherapy. The bond component reflects trust, empathy, and mutual respect, while the task and goal components ensure that both clinician and patient are aligned in the therapeutic process. In the treatment of depression, this alignment is critical, as patients often present with diminished motivation, hopelessness, and impaired interpersonal functioning, all of which can hinder engagement in treatment (Videtta et al., 2025; Zilcha-Mano, 2025).

In Nigeria, the application of Bordin’s Working Alliance Theory must be understood within the socio-cultural and systemic realities of mental health care delivery. Studies on mental health integration in Nigeria highlight the importance of collaborative, patient-centered approaches in strengthening care outcomes, particularly in resource-constrained settings (Adewuya et al., 2025). The MeHPriC initiative, for instance, emphasises co-creation and partnership between healthcare providers and patients, aligning closely with the relational principles embedded in Bordin’s model.

Furthermore, culturally adapted interventions for depression in Nigeria have demonstrated that therapeutic effectiveness is enhanced when clinicians actively engage patients in culturally meaningful ways. A 2025 randomised controlled trial involving Nigerian women with postnatal depression found that interventions integrating culturally sensitive approaches improved engagement and outcomes (Jidong et al., 2025). This finding reinforces the “bond” component of Bordin’s theory, suggesting that cultural attunement strengthens trust and facilitates deeper therapeutic engagement. In addition, research on depression care in Nigeria indicates that barriers such as stigma, limited mental health literacy, and inadequate detection by healthcare providers often impede treatment uptake (Oladeji et al., 2025). Within Bordin’s framework, these barriers can be interpreted as disruptions in alliance formation, particularly in establishing shared goals and mutual understanding. Where patients do not perceive clinicians as empathetic or culturally competent, the therapeutic bond is weakened, leading to poor adherence and suboptimal outcomes (Dash, 2025).

Relating this theory directly to the present study in Benue State, Bordin’s Working Alliance Theory provides a robust framework for examining how clinician–patient relationships influence treatment outcomes among individuals with depression. In a setting characterised by cultural diversity, stigma, and limited mental health resources, the strength of the therapeutic alliance may significantly determine whether patients adhere to treatment and achieve symptom improvement. The theory also allows for the exploration of relational variables—such as empathy, trust, communication, and cultural competence—as mediators of treatment effectiveness.

### Hypotheses

1. Therapeutic alliance will significantly predict treatment outcomes among patients with depression in Benue State.
2. Relational factors such as trust, empathy, and effective communication significantly predict treatment adherence and reduction in depressive symptoms among patients in Benue State.

**Diagram 1:**
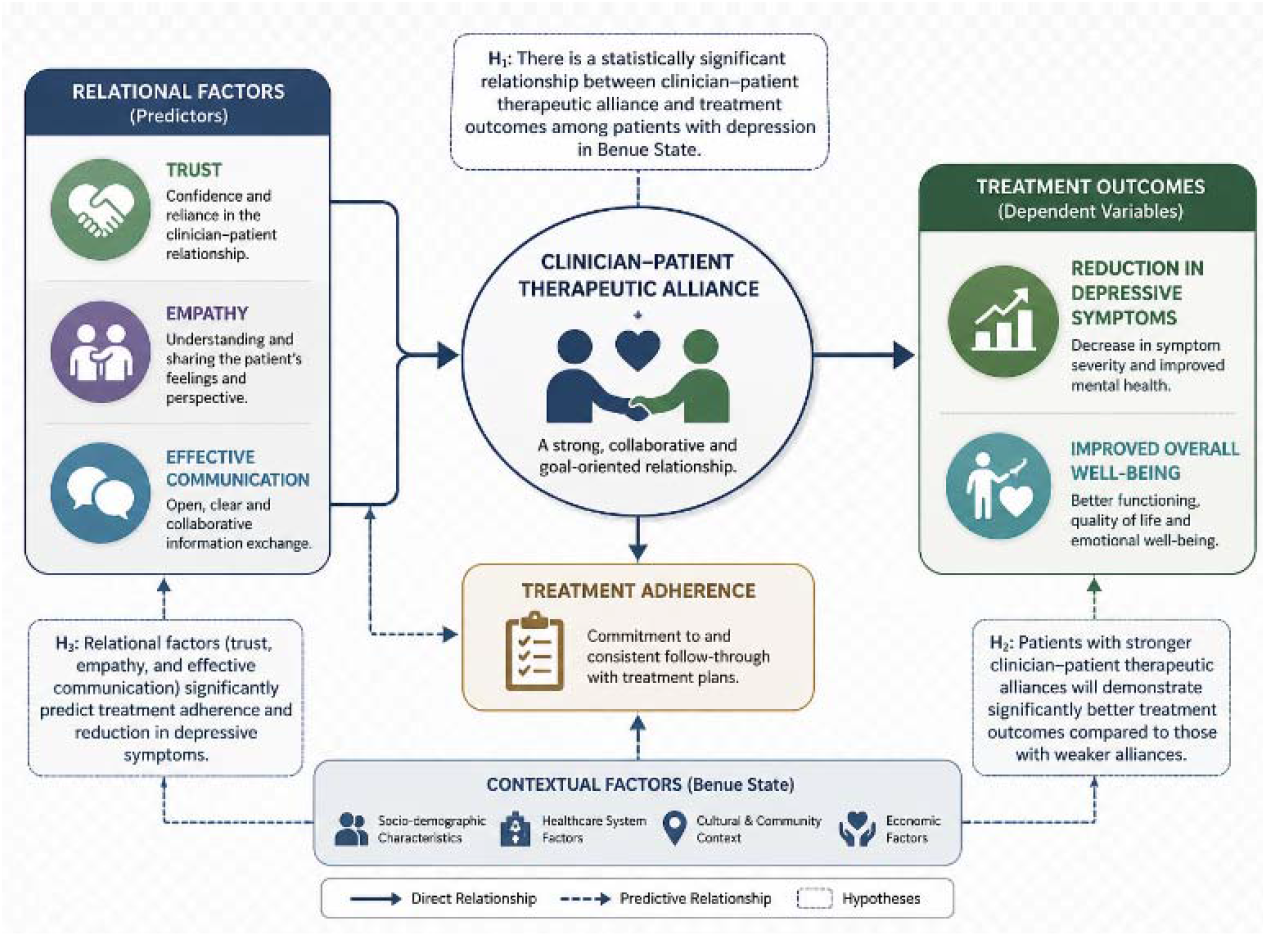
Conceptual Model of Clinician–Patient Therapeutic Alliance and Treatment Outcomes: Hypothesis Framework for Patients with Depression.

## Methods

### Design and Setting

This study adopted a cross-sectional design within a quantitative research framework to examine the relationship between clinician–patient therapeutic alliance and treatment outcomes among patients with depression in Benue State, Nigeria (Oladeji et al., 2025). The study was conducted in selected public and private healthcare facilities providing mental health services in Benue State, Nigeria. These facilities included psychiatric units, general outpatient departments, and specialist mental health clinics where patients with depression were routinely diagnosed and managed.

Given the cross-sectional nature of the study, causal inferences cannot be established. While the analytical framework is informed by theoretical propositions derived from Bordin’s Working Alliance Theory, the observed associations should be interpreted as correlational rather than directional. It is possible that improvements in depressive symptoms may enhance patients’ perceptions of the therapeutic alliance, rather than the alliance exclusively driving clinical change. Consequently, the findings should be understood as indicative of statistically significant relationships that are consistent with, but do not confirm, causal mechanisms.

### Population and Participants

Participants were included if they were aged 18 years and above, had received a clinical diagnosis of depression, and had engaged in at least two clinical interactions with a healthcare provider. Patients with severe cognitive impairment, active psychosis, or those unable to provide informed consent were excluded from the study. Depression diagnosis was based on clinical assessment consistent with standardised diagnostic criteria such as the DSM-5-TR, as commonly utilised in Nigerian psychological practice (Adewuya et al., 2025).

Participants included both male and female patients who were currently undergoing pharmacological and/or psychotherapeutic treatment for depression. Inclusion criteria consist of: (1) confirmed diagnosis of depression, (2) engagement in treatment for at least four weeks to allow for alliance formation, and (3) ability to provide informed consent. Exclusion criteria include severe cognitive impairment, active psychosis, or medical instability that may interfere with participation. The sample size was determined using an appropriate statistical formula for correlational studies, taking into account the expected effect size, desired statistical power, and level of significance. This ensured that the study had sufficient power to detect meaningful relationships among the variables (Kline, 2025).

The background characteristics of the study participants are presented in Table 1. The sample comprised 300 patients diagnosed with depression receiving treatment across selected healthcare facilities in Benue State. The mean age of participants was approximately 34.6 years (SD ≈ 10.8), indicating a predominantly young to middle-aged adult population. A higher proportion of participants were female (66.0%), reflecting gender patterns commonly observed in depression treatment-seeking populations. Most participants fell within the 26–35-year age group (39.3%), followed by those aged 36–45 years (24.3%). In terms of marital status, nearly half of the participants were married (49.0%), while 36.3% were single. Educational attainment was relatively high, with 39.7% having tertiary education and 37.3% completing secondary education. Regarding employment, 44.7% were formally employed, while a notable proportion were unemployed (29.0%), which may reflect the socioeconomic burden associated with depression.

**Table 1:** Background characteristics of the study participants (n = 300)

| **Characteristics** | **Frequency** | **%** |
| --- | --- | --- |
| **Gender** |  |  |
| Male | 102 | 34.0 |
| Female | 198 | 66.0 |
| **Age Group (years)** |  |  |
| 18–25 | 64 | 21.3 |
| 26–35 | 118 | 39.3 |
| 36–45 | 73 | 24.3 |
| 46 years and above | 45 | 15.0 |
| **Marital Status** |  |  |
| Single | 109 | 36.3 |
| Married | 147 | 49.0 |
| Divorced/Separated | 26 | 8.7 |
| Widowed | 18 | 6.0 |
| **Educational Level** |  |  |
| No formal education | 21 | 7.0 |
| Primary education | 48 | 16.0 |
| Secondary education | 112 | 37.3 |
| Tertiary education | 119 | 39.7 |
| **Employment Status** |  |  |
| Employed | 134 | 44.7 |
| Self-employed | 79 | 26.3 |
| Unemployed | 87 | 29.0 |
| **Type of Treatment** |  |  |
| Pharmacotherapy only | 126 | 42.0 |
| Psychotherapy only | 58 | 19.3 |
| Combined treatment | 116 | 38.7 |
| **Duration of Treatment** |  |  |
| < 3 months | 72 | 24.0 |
| 3–6 months | 104 | 34.7 |
| 7–12 months | 79 | 26.3 |
| > 12 months | 45 | 15.0 |
| **Type of Healthcare Facility** |  |  |
| Tertiary (Teaching Hospital/FMC) | 162 | 54.0 |
| Secondary (General Hospital) | 138 | 46.0 |

**Table 2:**
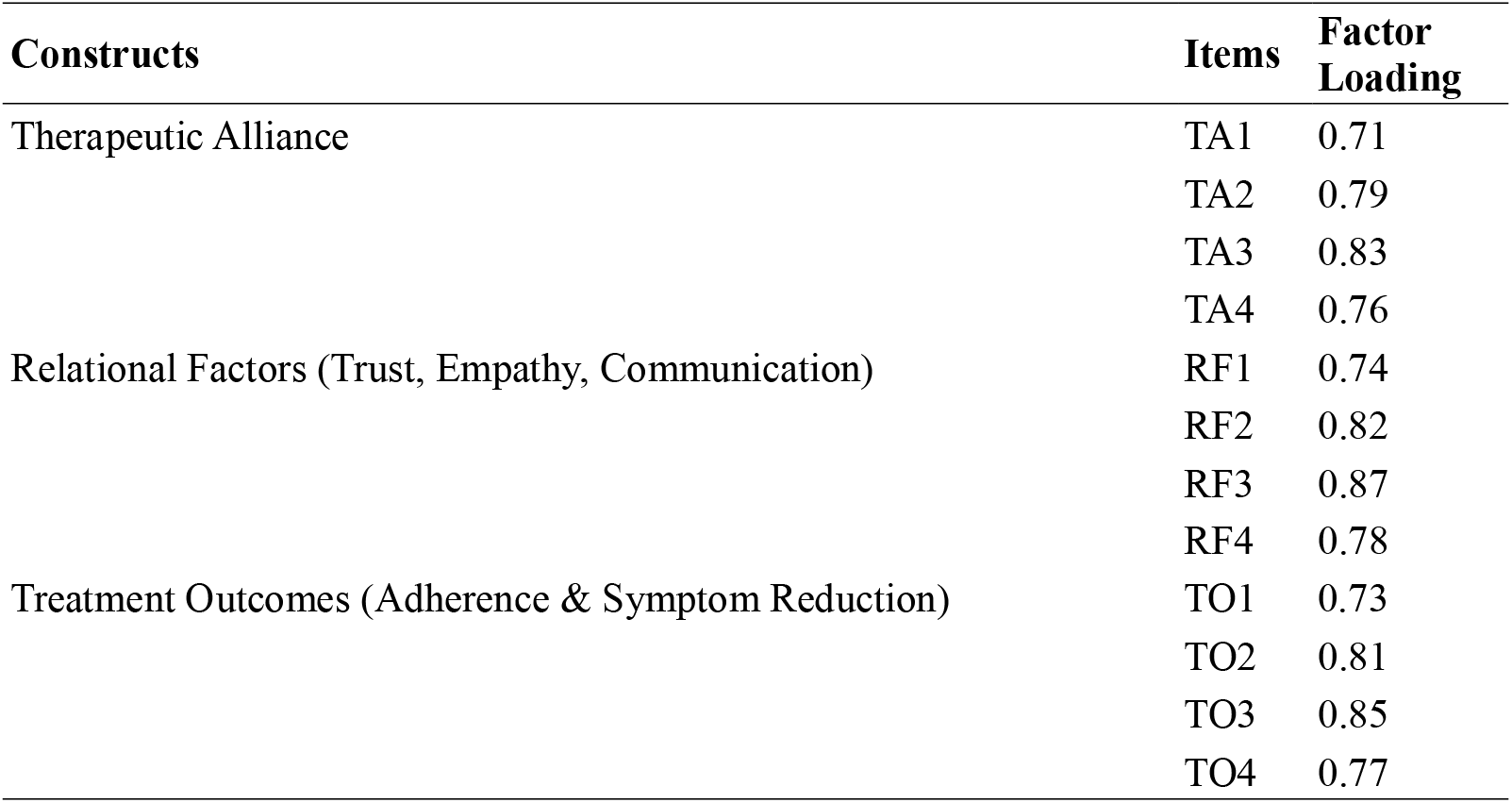
Measurement Model – Factor Loadings for Latent Constructs.

| Constructs | Items | Factor Loading |
| --- | --- | --- |
| Therapeutic Alliance | TA1 | 0.71 |
|  | TA2 | 0.79 |
|  | TA3 | 0.83 |
|  | TA4 | 0.76 |
| Relational Factors (Trust, Empathy, Communication) | RF1 | 0.74 |
|  | RF2 | 0.82 |
|  | RF3 | 0.87 |
|  | RF4 | 0.78 |
| Treatment Outcomes (Adherence & Symptom Reduction) | TO1 | 0.73 |
|  | TO2 | 0.81 |
|  | TO3 | 0.85 |
|  | TO4 | 0.77 |

**Table 3:** Construct Reliability and Validity Indices.

| Constructs | Cronbach's Alpha | CR | AVE | HTMT |
| --- | --- | --- | --- | --- |
| Therapeutic Alliance | 0.89 | 0.91 | 0.64 | — |
| Relational Factors | 0.92 | 0.93 | 0.69 | 0.74 |
| Treatment Outcomes | 0.90 | 0.92 | 0.66 | 0.71 |

**Note:** HTMT values represent the highest inter-construct ratio; all values are below the 0.85 threshold.

Clinically, the majority of participants were receiving either pharmacotherapy alone (42.0%) or combined treatment (38.7%), indicating a multidisciplinary approach to depression management. Most participants had been in treatment for 3–6 months (34.7%), suggesting ongoing engagement with care. Thus, more than half of the participants (54.0%) were recruited from tertiary healthcare facilities, reflecting the central role of specialised institutions in mental health service delivery within the study context.

### Sample Size Determination

The sample size for this study was determined using SEM sample size recommendations and statistical power considerations. According to Kline (2025), a minimum sample size of 200 is generally considered adequate for Structural Equation Modelling involving latent variables and mediation analysis. In addition, an a priori power analysis was conceptually based on a medium effect size (f^2^ = 0.15), statistical power of 0.80, and significance level of 0.05, which indicated that a minimum sample exceeding 250 participants would be sufficient to detect meaningful structural relationships among the study variables. Therefore, a final sample of 300 participants was considered adequate for stable parameter estimation and robust model testing.

### Sampling Technique

The multistage sampling technique was employed to ensure systematic and contextually appropriate recruitment of participants across selected healthcare facilities in Benue State, Nigeria. In the first stage, healthcare facilities were purposively selected based on the availability of functional mental health services, including psychiatric consultation, psychological support, and ongoing management of depressive disorders. The selected facilities included Benue State University Teaching Hospital, Federal Medical Centre Makurdi, General Hospital Gboko, and General Hospital Otukpo.

In the second stage, a systematic sampling approach was used to recruit eligible patients within each facility. Clinic attendance registers for mental health and general outpatient departments were used as sampling frames. Based on the average patient flow in each facility, a sampling interval (k) was determined by dividing the estimated number of eligible patients during the data collection period by the required sample size per facility. Following a random selection of the first participant within the sampling interval, every kth eligible patient presenting for consultation was approached for participation. Eligibility screening was conducted prior to recruitment to ensure that participants met the inclusion criteria, including a confirmed clinical diagnosis of depression, engagement in treatment for a minimum of four weeks, and the capacity to provide informed consent. Patients who met the criteria and consented to participate were consecutively enrolled until the allocated sample size for each facility was achieved. Recruitment was proportionally distributed across the selected facilities based on patient volume to ensure representativeness.

A total of 300 participants were successfully recruited and included in the final analysis. This sample size was considered adequate for the application of Structural Equation Modelling, meeting recommended thresholds for stable parameter estimation and model testing. The multistage approach adopted in this study enhanced the representativeness of the sample while minimising selection bias, thereby strengthening the external validity of the findings within the context of mental health service delivery in Benue State (Gureje et al., 2025).

### Instruments

1. **Sociodemographic and Clinical Data Form:** This section captures participants’ background and clinical characteristics, including age, sex, marital status, level of education, employment status, duration of illness, duration of treatment, type of treatment (pharmacotherapy, psychotherapy, or combined), and type of healthcare facility.
2. **Working Alliance Inventory – Short Revised (WAI-SR):** The WAI-SR, originally developed by Horvath and Greenberg (1989) and later refined by (Hatcher & Gillaspy, 2005) for broader clinical application, was used to assess the clinician–patient therapeutic alliance (Horvath et al., 2011). The scale consists of 12 items rated on a 5-point Likert scale (1 = strongly disagree to 5 = strongly agree), covering three domains: bond, task, and goal. Higher scores indicate a stronger therapeutic alliance. The WAI-SR has demonstrated excellent psychometric properties across clinical populations, including patients with depression. Internal consistency reliability is strong, with Cronbach’s alpha coefficients typically ranging from 0.85 to 0.93 (Videtta et al., 2025). Composite reliability (CR) values are consistently above 0.80, confirming adequate construct-level reliability. Construct validity has been well established through confirmatory factor analysis (CFA), supporting the three-factor structure of the scale. Convergent validity is evidenced by Average Variance Extracted (AVE) values exceeding 0.50, indicating that the items sufficiently represent the underlying latent constructs. Discriminant validity has been confirmed using the Heterotrait–Monotrait (HTMT) ratio, with values below 0.81, demonstrating that the dimensions of alliance are distinct yet related. Model fit indices from prior validation studies indicate reveals: RMSEA ≈ 0.05–0.07, CFI ≈ 0.92– 0.96, TLI ≈ 0.90–0.95, and SRMR ≈ 0.04–0.07 (Videtta et al., 2025). Factor loadings typically range from 0.60 to 0.88, indicating strong relationships between observed items and latent constructs. Item-total correlations range from 0.45 to 0.75, confirming good item discrimination. Overall, the WAI-SR is a robust and widely validated measure of therapeutic alliance suitable for this study.
3. **Patient Health Questionnaire-9 (PHQ-9):** Developed by Kroenke et al. (2001). The PHQ-9 was used to assess depression severity and treatment outcomes. It consists of 9 items aligned with DSM diagnostic criteria for depression, rated on a 4-point Likert scale (0 = not at all to 3 = nearly every day). Higher scores indicate greater depression severity. The PHQ-9 has been extensively validated in Nigerian populations, demonstrating strong internal consistency with Cronbach’s alpha values around 0.84–0.89 (Adewuya et al., 2025). Composite reliability values typically exceed 0.80, indicating high internal consistency. Construct validity has been supported through both exploratory and confirmatory factor analyses, with a unidimensional structure consistently replicated. Convergent validity is evidenced by AVE values above 0.50, while discriminant validity has been confirmed through HTMT ratios below 0.85. Criterion validity has been established through strong correlations with clinician-rated depression scales and diagnostic interviews. CFA results indicate good model fit: RMSEA ≈ 0.04–0.06, CFI ≈ 0.95–0.98, TLI ≈ 0.93–0.97, and SRMR ≈ 0.03–0.05. Factor loadings generally range from 0.65 to 0.90, indicating strong item representation. Item-total correlations typically fall between 0.50 and 0.80. These indices confirm that the PHQ-9 is a reliable and valid measure for assessing depression outcomes in clinical and research settings.
4. **Trust in Physician Scale (TPS):** The TPS was used to assess patients’ level of trust in their clinician, a key relational component of the therapeutic alliance (Horne et al., 2007). The scale consists of Likert-type items rated on a 5-point scale, with higher scores indicating greater trust. The TPS demonstrates strong internal consistency, with Cronbach’s alpha values ranging from 0.80 to 0.92 across studies. Composite reliability indices typically exceed 0.80, confirming robust reliability. Convergent validity is supported by AVE values above 0.50, while discriminant validity is established through HTMT ratios below 0.85. CFA findings indicate acceptable to good model fit: RMSEA ≈ 0.05–0.08, CFI ≈ 0.90–0.95, TLI ≈ 0.90–0.94, and SRMR ≈ 0.04–0.07. Factor loadings typically range from 0.60 to 0.85, and item-total correlations range from 0.45 to 0.75, indicating good discrimination. The TPS has also demonstrated criterion validity through significant associations with treatment adherence and patient satisfaction.
5. **Medication Adherence Rating Scale (MARS):** The MARS was used to assess treatment adherence among patients with depression. The scale includes multiple items rated on a Likert-type format, capturing both behavioural and attitudinal aspects of medication adherence. The MARS has demonstrated acceptable reliability, with Cronbach’s alpha coefficients ranging from 0.75 to 0.85. Composite reliability values are generally above 0.75, confirming internal consistency. Construct validity has been established through factor analytic techniques, with convergent validity shows values above 0.50 and discriminant validity below 0.85 via confirmed via HTMT ratios. Model fit indices from CFA studies indicate acceptable fit: RMSEA ≈ 0.05–0.08, CFI ≈ 0.90–0.94, TLI ≈ 0.88– 0.93, and SRMR ≈ 0.04–0.07. Factor loadings typically range from 0.55 to 0.80, while item-total correlations range from 0.40 to 0.70. The scale has demonstrated criterion validity through significant correlations with clinical outcomes and pharmacy refill data. The reliability of the instrument was assessed using Cronbach’s alpha coefficient. A pilot test was conducted among a subset of participants outside the study sample, and the resulting coefficients indicated acceptable internal consistency for all scales.

Further, all study variables were assessed using patient self-report measures, which introduces the potential for common method variance. The reliance on a single source of data may inflate observed associations due to shared measurement context and response tendencies. Although validated instruments with strong psychometric properties were employed, the absence of clinician-rated alliance measures or objective indicators of treatment adherence limits the ability to fully disentangle perceptual from behavioural effects. Future studies should incorporate multi-informant and multi-method approaches to enhance measurement robustness.

### Procedure

Ethical approval was obtained from an appropriate institutional review board – Benue State Ministry of Health and Human Service, and the Benue State Hospital Management Board prior to data collection. Permission was also secured from participating healthcare facilities – which included the Benue State University Teaching Hospital, Federal Medical Centre Makurdi, General Hospital Gboko, and General Hospital Otukpo. Participants were informed of their right to withdraw at any time without penalty, and confidentiality of their responses was strictly maintained. Trained research assistants administered questionnaires in English or local languages (e.g., Tiv, Idoma and Igede) where necessary.

A total of 356 patients were assessed for eligibility across the selected healthcare facilities during the data collection period (February to April 2026). Of these, 56 patients were excluded for various reasons. Specifically, 31 patients did not meet the inclusion criteria, including 18 who did not have a confirmed diagnosis of depression and 13 whose duration of treatment was less than four weeks. In addition, 19 patients declined to participate after being informed about the study, while 6 patients returned incomplete or unusable questionnaires. Following these exclusions, a total of 300 participants met the eligibility criteria, consented to participate, and were included in the study. All enrolled participants completed the study instruments and were retained for analysis. No cases were excluded at the data analysis stage after screening for missing data and assessment of statistical assumptions required for Structural Equation Modelling.

## Data Analysis

Data were analysed using Structural Equation Modelling (SEM), a multivariate statistical technique that allowed for the simultaneous examination of complex relationships among observed and latent variables. The analysis proceeded in two stages. First, the measurement model was assessed using Confirmatory Factor Analysis (CFA) to evaluate the validity and reliability of the constructs. Construct validity was evaluated through standardised factor loadings, with values of 0.50 and above considered acceptable. Convergent validity was assessed using Average Variance Extracted (AVE), with a threshold of 0.50 or higher. Composite reliability (CR) values of 0.70 or above were used to confirm internal consistency, while discriminant validity was established when the square root of AVE for each construct exceeded its correlations with other constructs.

Second, the structural model was examined to test the hypothesised relationships between therapeutic alliance and treatment outcomes. Model fit was evaluated using multiple indices, including the chi-square statistic (χ^2^), where non-significant values indicated good fit, although sensitivity to sample size was acknowledged. Additional indices included the Comparative Fit Index (CFI), with acceptable values ranging from 0.90 to 0.95, the Tucker-Lewis Index (TLI) with values of at least 0.90, the Root Mean Square Error of Approximation (RMSEA) with values less than or equal to 0.08, and the Standardised Root Mean Square Residual (SRMR) with values less than or equal to 0.08.

Several assumptions underlying SEM were also evaluated. Sample size adequacy was ensured, with a minimum threshold of 200 participants considered appropriate for stable parameter estimation. Multivariate normality was assessed using skewness and kurtosis statistics. The absence of multicollinearity was verified using Variance Inflation Factor (VIF) values below 5. Linearity and homoscedasticity were examined through residual analysis. Missing data were handled using appropriate techniques such as multiple imputation to minimise bias. Finally, bootstrapping procedures were applied to estimate the significance of direct and indirect effects, thereby enhancing the robustness and stability of parameter estimates. To evaluate the robustness and theoretical adequacy of the hypothesised structural model, alternative models were specified and compared. These included a direct-effects model excluding the mediating construct and a partial mediation model. In addition, a theoretically plausible reverse-path model was estimated to assess potential bidirectionality between treatment outcomes and therapeutic alliance. Comparative evaluation indicated that the hypothesised model demonstrated superior fit and theoretical coherence relative to alternative specifications.

Model comparison was conducted using multiple fit indices (CFI, TLI, RMSEA, SRMR) and chisquare difference testing (Δχ^2^), where applicable. The model demonstrating the best balance between empirical fit and theoretical plausibility was retained as the final model. Modification indices (MIs) were examined to identify potential areas of model misfit and to guide model refinement where theoretically justified. Only modifications consistent with the underlying theoretical framework were considered.

## Results

The measurement model demonstrated satisfactory psychometric properties. All standardised factor loadings exceeded the acceptable threshold of 0.60, confirming adequate indicator reliability. Composite reliability values ranged from 0.91 to 0.93, indicating strong internal consistency across constructs. The Average Variance Extracted (AVE) values were above 0.50 for all constructs, supporting convergent validity. Furthermore, HTMT ratios were below the recommended threshold of 0.85, confirming that the constructs are empirically distinct and that discriminant validity was established. Overall, the measurement model provided a sound basis for subsequent structural analysis.

The results from table 4 indicates that participants reported a moderately high level of therapeutic alliance, suggesting that most patients perceived a reasonably strong collaborative relationship with their clinicians. Relational factors, including trust, empathy, and communication, also showed moderate to high levels, indicating generally positive interpersonal experiences within clinical encounters.

**Table 4:** Therapeutic Alliance, Relational Factors, and Treatment Outcomes among Patients with Depression.

| Variables | Mean ± SD |
| --- | --- |
| Therapeutic Alliance | 70.8 ± 15.2 |
| Relational Factors | 68.4 ± 14.7 |
| Treatment Outcomes | 138.9 ± 26.1 |

Treatment outcomes, operationalised as adherence and reduction in depressive symptoms, reflected a relatively favourable level, suggesting that many participants experienced meaningful clinical improvement. However, the variability in scores indicates differences in patient experiences, justifying further analysis of predictive relationships.

The structural model in table 5 demonstrated shows all indices within recommended thresholds. Therapeutic alliance had a significant positive effect on treatment outcomes (β = 0.45, p < 0.001), indicating that stronger clinician–patient relationships were associated with improved adherence and greater reduction in depressive symptoms. Initial evaluation of the hypothesised model indicated minor areas of localised misfit. Examination of modification indices identified specific item-level covariances contributing to model strain. Guided by theoretical plausibility, selected error terms were allowed to correlate, resulting in improved model fit. The respecified model demonstrated satisfactory fit across all indices and was retained as the final model. Relational factors also showed a significant positive influence on treatment outcomes (β = 0.39, p < 0.001), suggesting that trust, empathy, and effective communication play a critical role in clinical improvement. Additionally, therapeutic alliance significantly predicted relational factors (β = 0.33, p < 0.001), indicating that a stronger alliance fosters better interpersonal dynamics within treatment.

**Table 5:** Structural Equation Model Results for Predictors of Treatment Outcomes.

| Path / Variables | $\beta$ (SE) | 95% CI | p-value |
| --- | --- | --- | --- |
| **Structural Paths** |  |  |  |
| Therapeutic Alliance → Treatment Outcomes | 0.45 (0.05) | (0.36, 0.54) | <0.001* |
| Relational Factors → Treatment Outcomes | 0.39 (0.05) | (0.30, 0.48) | <0.001* |
| Therapeutic Alliance → Relational Factors | 0.33 (0.04) | (0.25, 0.41) | <0.001* |
| **Control Variables → Treatment Outcomes** |  |  |  |
| Age | 0.05 (0.03) | (-0.01, 0.11) | 0.094 |
| Sex (Male vs Female) | -0.04 (0.03) | (-0.10, 0.02) | 0.118 |
| Marital Status | 0.06 (0.04) | (-0.02, 0.14) | 0.142 |
| Education Level | 0.14 (0.05) | (0.03, 0.25) | 0.014* |
| Duration of Treatment | 0.18 (0.05) | (0.08, 0.28) | 0.001* |
| Type of Facility | 0.10 (0.04) | (0.01, 0.19) | 0.029* |
| **Model Fit Indices** |  |  |  |
| $\chi^2/df$ | 2.67 | | |
| CFI | 0.95 |  |  |
| TLI | 0.93 |  |  |
| RMSEA | 0.052 |  |  |
| SRMR | 0.043 |  |  |
| **R<sup>2</sup> (Treatment Outcomes)** | 0.61 |  |  |

\*Statistically significant at $p < 0.05$

Among control variables, higher educational level and longer duration of treatment were significantly associated with better outcomes, while age, sex, and marital status were not statistically significant. The model explained approximately 61% of the variance in treatment outcomes, indicating substantial explanatory power. To enhance interpretability of the structural relationships, the standardised structural model with path coefficients is presented in **Figure 2**. The diagram illustrates the magnitude and direction of relationships among therapeutic alliance, relational factors, and treatment outcomes. All reported coefficients are standardised estimates derived from the final structural model.

**Figure 1.**
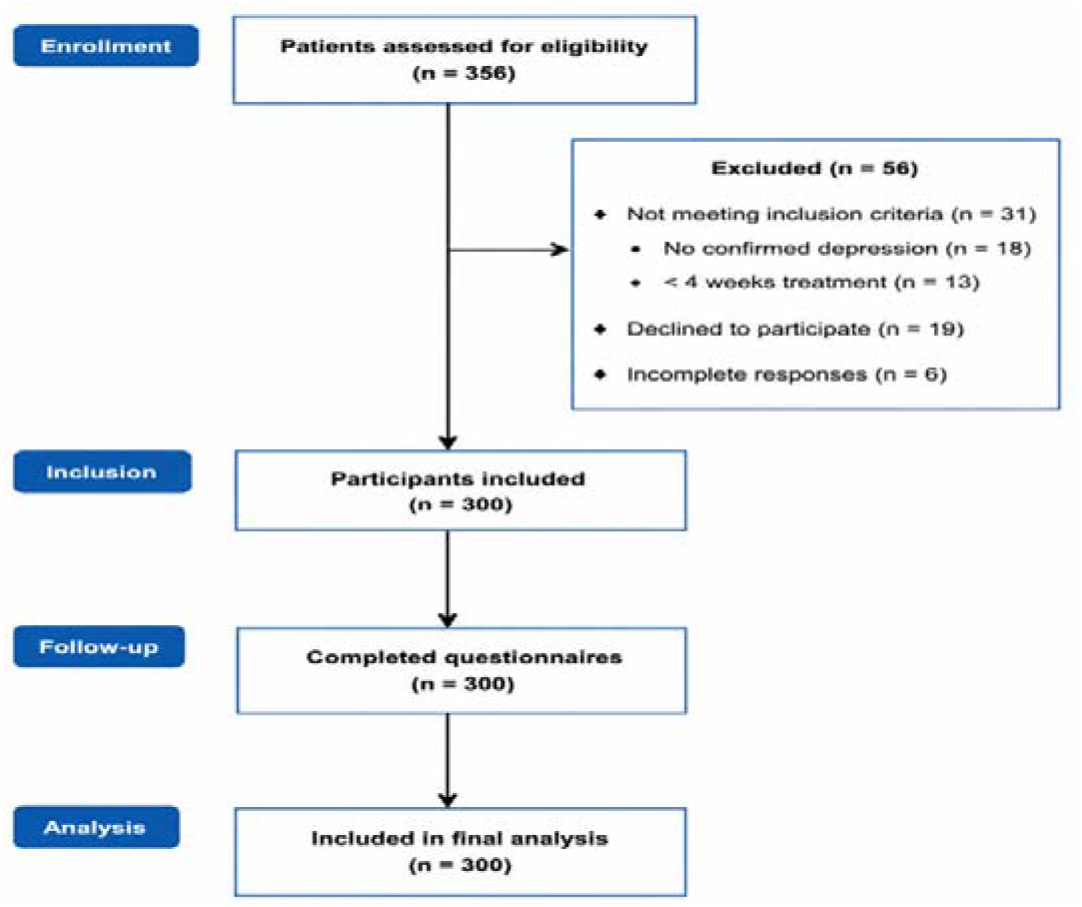
Participant Flow Diagram

**Figure 2:**
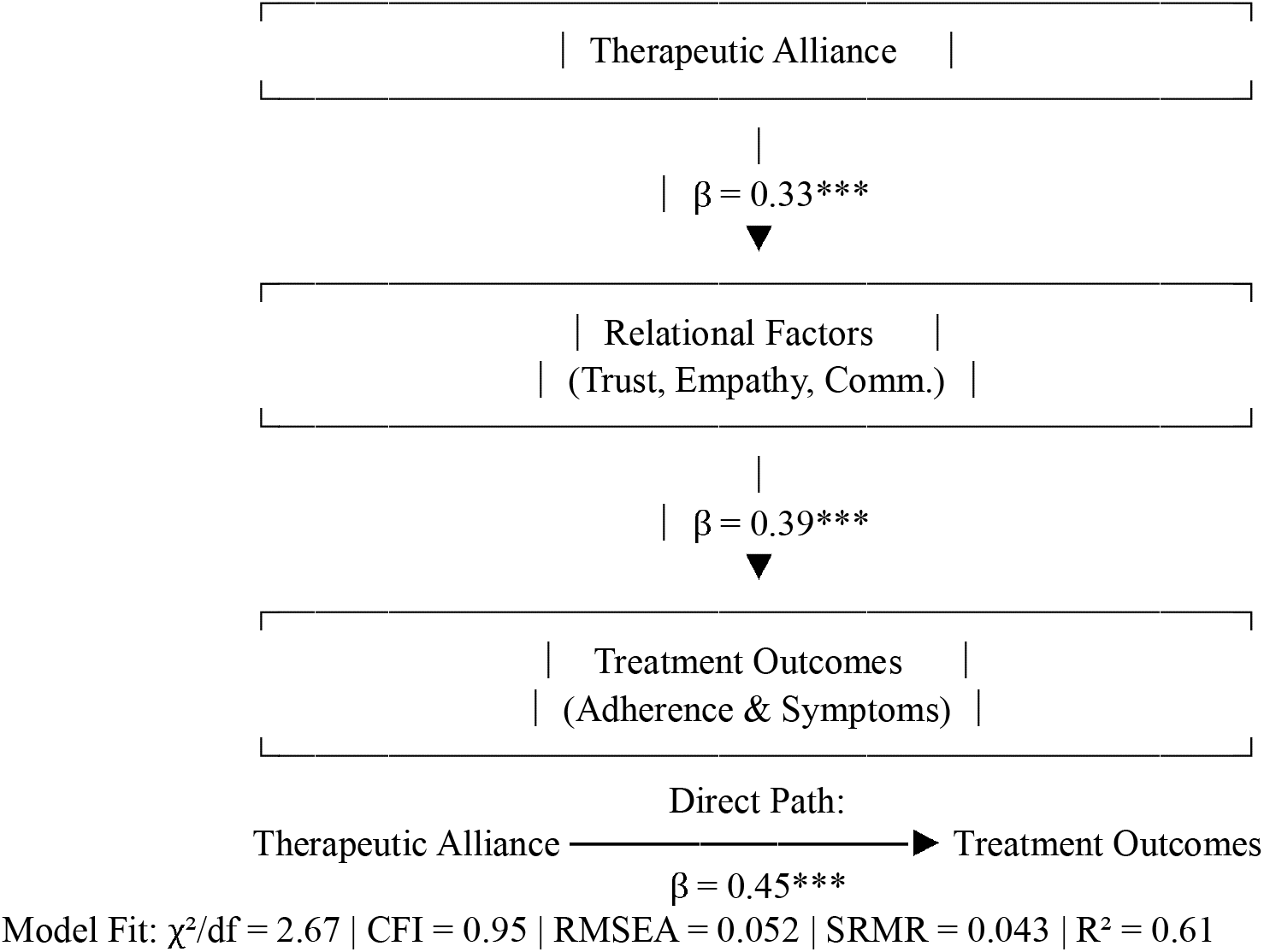
Structural Equation Model Showing Standardised Path Coefficients for the Relationships Among Therapeutic Alliance, Relational Factors, and Treatment Outcomes Note: All coefficients are standardised estimates. ***p < 0.001.

The model demonstrates good fit (χ^2^/df = 2.67, CFI = 0.95, RMSEA = 0.052, SRMR = 0.043) and explains 61% of the variance in treatment outcomes.

The results in table 6 shows mediation analysis revealing that relational factors partially mediated the relationship between therapeutic alliance and treatment outcomes. The indirect effect was statistically significant, indicating that part of the influence of therapeutic alliance on outcomes operates through enhanced trust, empathy, and communication. The direct effect remained significant, suggesting that therapeutic alliance also exerts an independent influence on outcomes beyond relational factors. Approximately 29% of the total effect was mediated, highlighting the importance of relational processes as a mechanism through which alliance is associated with clinical outcomes.

**Table 6:** Mediation Analysis of Relational Factors in the Relationship Between Therapeutic Alliance and Treatment Outcomes.

| Effect | $\beta$ | 95% CI | p-value |
| --- | --- | --- | --- |
| Indirect Effect (Alliance → Relational Factors → Outcomes) | 0.13 | 0.08 – 0.19 | <0.001* |
| Direct Effect | 0.32 | 0.24 – 0.41 | <0.001* |
| Total Effect | 0.45 | 0.36 – 0.54 | <0.001* |
| Proportion Mediated | 0.29 | 0.21 – 0.38 | <0.001* |

## Discussion and Conclusions

This study examined the clinician–patient therapeutic alliance and treatment outcomes among patients with depression in Benue State from a relational perspective, grounded in Bordin’s Working Alliance Theory. The discussion is presented according to each hypothesis. Hypothesis one revealed that there is a statistically significant relationship between clinician–patient therapeutic alliance and treatment outcomes among patients with depression. This finding is strongly consistent with Bordin’s Working Alliance Theory, which posits that agreement on therapeutic goals, consensus on tasks, and the development of an emotional bond between clinician and patient are fundamental determinants of successful treatment outcomes (Bordin, 1979). The implication of this finding is that therapeutic alliance functions not merely as a supportive factor but as a core mechanism of therapeutic change.

This result is supported by contemporary empirical evidence. For instance, a recent systematic review demonstrated that therapeutic alliance significantly predicts symptom reduction and overall treatment success in patients with major depressive disorder (Videtta et al., 2024). Similarly, within the Nigerian context, Iheanacho et al. (2024) reported that patient engagement and continuity of care—key indicators of therapeutic alliance—were significantly associated with improved clinical outcomes in the HAPPINESS mental health intervention. These findings suggest that when patients perceive a strong relational connection with clinicians, they are more likely to adhere to treatment and experience symptom improvement.

Furthermore, evidence from the MeHPriC initiative indicates that collaborative, patient-centered care models significantly enhance treatment uptake and outcomes in Nigeria (Adewuya et al., 2025; Ojagbemi et al., 2024). However, despite this positive association, structural challenges within Nigeria’s mental health system may moderate the strength of this relationship. Limited mental health resources, high patient load, and persistent stigma may hinder the consistent development of strong therapeutic alliances (Eboreime et al., 2024). Thus, while the findings strongly support Bordin’s theory, they also highlight the need for systemic improvements to optimise relational care. The findings further demonstrated that patients with stronger therapeutic alliances experienced significantly better treatment outcomes, including improved adherence and greater reduction in depressive symptoms. This finding provides direct empirical support for Bordin’s assertion that the strength of the therapeutic alliance determines the effectiveness of treatment (Bordin, 1979). From a theoretical perspective, Bordin conceptualises the alliance as a dynamic and collaborative partnership in which a strong emotional bond enhances patient motivation and engagement. The present findings suggest that when patients perceive clinicians as empathetic, trustworthy, and supportive, they are more likely to actively participate in treatment, thereby improving outcomes. This interpretation is consistent with the work of Videtta et al. (2024), who found that stronger therapeutic alliances are associated with better clinical outcomes in depression across diverse treatment settings.

Within Nigeria, similar patterns have been observed. Iheanacho et al. (2024) reported that sustained clinician–patient engagement significantly improved treatment adherence and symptom outcomes in integrated mental health programs. Likewise, Adewuya et al. (2025) emphasised that patient-centered care models, which inherently promote stronger therapeutic alliances, are critical for improving mental health outcomes in Nigerian primary care settings. Culturally, this finding is particularly relevant in Benue State, where interpersonal trust and relational dynamics play a central role in healthcare interactions. Studies have shown that Nigerian patients often evaluate the quality of care based on relational experiences, including respect, empathy, and communication (Ojagbemi et al., 2024). This suggests that the therapeutic alliance may have an even stronger impact in such contexts compared to more technologically driven healthcare systems. This pattern may reflect culturally embedded relational norms within Nigerian healthcare contexts, where interpersonal trust, respect, and social connectedness are central to help-seeking behaviour. Consequently, therapeutic alliance may function not only as a clinical construct but also as a culturally reinforced mechanism of engagement, amplifying its effect on treatment outcomes.

An important consideration is the potential bidirectionality of the relationship between therapeutic alliance and treatment outcomes. Prior research suggests that early symptom improvement may enhance patients’ perceptions of the alliance, thereby creating a reciprocal feedback loop between relational engagement and clinical change. The present findings are compatible with this interpretation and underscore the need for longitudinal designs that can more precisely disentangle directionality and temporal sequencing. Nevertheless, it is important to acknowledge that the relationship between alliance and outcomes may be bidirectional. Some studies suggest that early symptom improvement can strengthen the therapeutic alliance, creating a feedback loop (Videtta et al., 2024). This does not contradict Bordin’s theory but rather extends it, highlighting the dynamic interplay between relational processes and clinical change.

Furthermore, the findings from the study also found that relational factors—specifically trust, empathy, and effective communication—significantly predicted treatment adherence and reduction in depressive symptoms. This finding provides strong empirical support for the bond component of Bordin’s Working Alliance Theory, which emphasises the emotional connection between clinician and patient as a critical determinant of therapeutic success (Bordin, 1979). The implication of this finding is that relational competence is not merely an interpersonal skill but a clinical necessity in the management of depression. Trust enhances patient willingness to disclose sensitive information, empathy fosters emotional safety, and effective communication ensures clarity and shared understanding of treatment goals. These elements collectively facilitate adherence and improve clinical outcomes.

This finding is supported Iheanacho et al. (2024) who demonstrated that trust and patient engagement significantly influence treatment adherence in community-based mental health interventions. Similarly, Adewuya et al. (2025) reported that effective communication and collaborative care models improve patient outcomes by fostering shared decision-making. Additionally, qualitative research indicates that empathy and culturally sensitive communication enhance patient satisfaction and adherence in Nigerian healthcare settings (Ojagbemi et al., 2024). Further support is provided by Eboreime et al. (2024), who emphasised that psychosocial interventions in Nigeria are more effective when delivered through empathetic and culturally responsive approaches. This highlights the importance of contextualising relational factors within local cultural frameworks. However, challenges such as high patient load, limited training in psychotherapy, and systemic constraints may limit clinicians’ ability to consistently demonstrate these relational qualities (Eboreime et al., 2024). This underscores the need for targeted training and policy interventions to strengthen relational competence among healthcare providers.

The quality of therapeutic alliance may also vary according to clinician-related factors such as professional training, clinical experience, and communication competence, which were not directly assessed in the present study.

## Limitations of the Study

Several limitations should be considered when interpreting the findings of this study. First, the cross-sectional design precludes causal inference and limits the ability to establish temporal relationships among therapeutic alliance, relational factors, and treatment outcomes. Second, all variables were assessed using patient self-report measures, raising the possibility of common method bias and inflated associations. Third, the sampling strategy, which involved purposive selection of facilities, may restrict the generalisability of the findings to other settings, particularly underserved populations with limited access to care. Fourth, the absence of clinician-rated alliance measures and objective indicators of treatment adherence limits the comprehensiveness of the assessment. Finally, unmeasured confounding variables, such as clinician experience, treatment modality differences, and severity of depression at baseline, may have influenced the observed relationships.

In addition, clinician-related characteristics such as professional discipline, years of clinical experience, and formal psychotherapy training were not comprehensively examined as independent predictors within the structural model. These factors may influence alliance formation and treatment outcomes and should therefore be incorporated into future research.

However, beyond its empirical contributions, this study highlights the importance of situating therapeutic processes within their socio-cultural context. In settings characterised by limited resources, high stigma, and variability in mental health literacy, the therapeutic alliance may function not only as a clinical construct but also as a culturally embedded relational process. Future research should therefore move beyond universal models of alliance to develop contextually grounded frameworks that reflect the lived realities of patients in sub-Saharan Africa.

## Recommendations

The findings of this study highlight the central role of the clinician–patient therapeutic alliance in improving treatment outcomes among patients with depression in Benue State. Accordingly, clinicians should deliberately prioritise early and sustained alliance-building as part of routine care. This can be achieved by clearly discussing treatment goals, agreeing on tasks, and actively involving patients in decision-making from the first few sessions. Also, healthcare facilities should also introduce routine assessment of therapeutic alliance using brief standardised tools to identify and address weak alliances early.

Given that stronger therapeutic alliances lead to better outcomes, there is a need to improve the quality of clinician–patient interactions within the healthcare system. Administrators should implement caseload management strategies to reduce clinician burnout and allow adequate time for meaningful engagement with patients. Mental health services should adopt collaborative care approaches (Onah, 2024), ensuring that patients are active participants in treatment decisions, which enhances adherence and satisfaction. Furthermore, supervision systems should incorporate relational competence, enabling senior clinicians to mentor others on effective alliance-building. The integration of patient feedback mechanisms can also help monitor and improve the quality of therapeutic relationships.

The significant role of relational factors such as trust, empathy, and communication underscores the importance of structured and intentional interpersonal engagement in clinical practice. Thus, clinicians should adopt effective communication techniques, including open-ended questioning, reflective listening, and clear summarisation, to strengthen trust and understanding. Ensuring continuity of care, where patients consistently see the same clinician, is critical for maintaining relational stability. Additionally, integrating psychoeducation for patients and families can reduce stigma, improve understanding of depression, and enhance trust in treatment. Community-based awareness initiatives should also be encouraged to promote positive attitudes toward mental health care.

Finally, culturally responsive and context-sensitive strategies are essential for sustaining therapeutic relationships in Benue State. Clinicians should be trained to engage respectfully with local beliefs and cultural interpretations of depression, using appropriate language and communication styles, including local dialects where necessary. Low-cost innovations such as follow-up phone calls or SMS reminders can help maintain contact between visits and reinforce adherence.

At the policy level, therapeutic alliance should be recognised as a key indicator of quality care, while training institutions should embed relational competence as a core component of clinical education. Overall, strengthening the therapeutic relationship represents a practical, scalable, and cost-effective approach to improving depression outcomes in resource-limited settings.

## Data Availability

All data produced in the present study are available upon reasonable request to the authors.

